# Minimal Role of Hamstring Hypertrophy in Strength Transfer Between Nordic Hamstring and Stiff-Leg Deadlift: A Blinded Randomized Controlled Trial

**DOI:** 10.1101/2024.12.17.24319207

**Authors:** Titouan Morin, Valentin Doguet, Antoine Nordez, Arnault Caillet, Lilian Lacourpaille

## Abstract

Strength transfer refers to the gain in strength in an untrained exercise resulting from training another exercise. This study aims to determine whether whole and selective hamstring hypertrophy influence the magnitude of strength transfer between the Nordic hamstring exercise (NHE) and the stiff-leg Deadlift (SDL). Using a blinded randomized controlled design, thirty-six resistance-untrained individuals were assigned either to a control group (CON), a NHE group, and a SDL group, the two resistance training programs being performed three times per week for nine weeks. Semimembranosus (SM), semitendinosus (ST), and biceps femoris (BF) hypertrophy was estimated from changes in the muscle volume. Strength transfer was measured by changes in the one-repetition maximum (1RM) of the non-trained exercise. After the resistance training programs, similar and significant whole hypertrophy was observed in both training groups (NHE: 11.4±6.5%, and SDL: 7.0±8.1%). The NHE group and SDL group, however, presented a selective hypertrophy of ST (24.3±10.8%) and SM (11.2±12.7%), respectively. Conversely, no difference in strength transfer was observed between the NHE group (10.7±8.5%) and the SDL group (20.7±15.0%) (p=0.06). Finally, non-significant correlations were found between strength transfer and both whole and selective hamstring hypertrophy (r≤0.3), except for the BF in SDL (r=0.6). We conclude that hamstring hypertrophy has a minimal role in strength transfer in resistance-untrained individuals. These findings suggest that, even in presence of hypertrophy, prevention and rehabilitation programs should include both Nordic hamstring and stiff-leg Deadlift exercises in the same training block to increase hamstring strength across both knee flexion-oriented and hip extension-oriented tasks.

## INTRODUCTION

The concept of strength transfer refers to the capacity of one exercise to improve strength in other, non-trained exercises or movement tasks (for review, see Mang et al. (23)). For instance, if a lifter performs kettlebell swings during a training block without performing a Deadlift, yet their Deadlift 1 repetition maximal (1RM) improves as a result of the kettlebell swings, this demonstrates a strength transfer between the two exercises (23). Strength transfer becomes then valuable since it allows strength and conditioning specialists to diversify their resistance training (RT) programs by incorporating exercises that yield additional and/or complementary effects.

Strength transfer magnitude seems to be influenced by several factors such as cross-task similarities in the external force vector direction (10,31), force-velocity constraints (20,32), and the number of degrees of freedom (e.g., mono- vs. bi-articular; unilateral vs. bilateral). Additionally, a common practical strategy lies in the use of “accessory exercises” to induce strength gains via hypertrophic increases of agonist muscles group while reducing stresses on muscular and non-muscular structures involved in the main exercise. This strategy is reasonable, as a hypertrophy-related increase in force-generating capacity of agonist muscles may lead to a greater force production, even for non-trained exercises. However, little is known about the role of muscle hypertrophy in strength transfer. One recent study showed that strength transfer to a three-repetition maximum (3RM) Deadlift was not different between groups that performed hip thrust (15%) and back squat training (16%), while similar gluteus maximus hypertrophy (11%) was induced in both groups (28). However, Carmichael et al. (8) reported that a comparable 16% increase in hamstring hypertrophy, achieved through a 6-week protocol of either eccentric or isometric hip extension exercises, did not lead to strength transfer during isometric or eccentric knee flexion, respectively. Strength transfer to knee flexion performance was observed only when the contraction regimen during training matched that of the testing conditions, i.e., eccentric and isometric hip extension training increased about 10.2% and 10.3% eccentric and isometric maximal knee flexion, respectively. Therefore, it remains uncertain whether hypertrophy directly contributes to strength transfer mechanisms.

Bi-articular hamstring muscle group, acting on both hip and knee joint, is an ideal model to understand the relationship between hypertrophy and strength transfer. This is because similar hypertrophy can be induced while using a mono-articular knee flexion-oriented and a hip extension-oriented movement training (5). Importantly, the distribution of hypertrophy among hamstring heads varies depending on the joint being mobilized. For instance, 10 weeks of Nordic hamstring training has been reported to induce preferential hypertrophy in the semitendinosus (5), whereas 10 weeks of stiff-leg Deadlift training would rather induce a preferential hypertrophy in the semimembranosus (18). These two individual muscles exhibited large anatomical differences (19), with a larger physiological cross section area for the SM leading to a greater torque generating capacities in knee flexion (113.3 u.a.) compared to ST (60.2 u.a.) (4). Therefore, if strength transfer depends on hypertrophy, stiff-leg Deadlift training that would rather hypertrophy a “stronger” muscle such as SM could exhibit greater strength transfer than Nordic hamstring training, more specific to a “weaker” ST muscle.

This study aimed to determine whether whole and selective hamstring hypertrophy influence the magnitude of strength transfer between two mono-articular movements: the knee flexion-oriented Nordic hamstring curl and the hip extension-oriented stiff-leg Deadlift. First, we hypothesized that individuals involved in stiff-leg Deadlift (SDL group) and Nordic hamstring (NHE group) resistance programs would demonstrate significant whole hypertrophy and strength transfer (5) compared to a control group (CON) not engaged in resistance training.

Second, we hypothesized that SDL group would show greater selective hypertrophy in SM, while the NHE group would exhibit greater hypertrophy in ST (5,18). Based on the SM’s higher torque generating capacity (4), we hypothesized that SM hypertrophy in the SDL group would result in a greater strength transfer (in Nordic hamstring exercise) than in the NHE group. For each group (NHE and SDL), we hypothesized that strength transfer would correlate significantly with both whole-hamstring and selective-hamstring hypertrophy.

To further investigate these mechanisms, surface EMG was used to measure individual hamstring muscle activation before and after training, providing insights to neural adaptations. These findings aim to enhance our understanding of the mechanisms underpinning strength transfer.

## METHODS

### Experimental Approach to the Problem

A randomized controlled trial (RCT) investigated the impact of hamstring muscle hypertrophy and its distribution across hamstring muscles on strength transfer between two exercises: the Nordic hamstring exercise and the stiff-leg Deadlift. The experimental protocol included three experimental sessions: a familiarization session and two testing sessions, which were conducted before (i.e., *pre*) and after (i.e., *post*) the resistance training program, which comprised 27 training sessions. Hamstring muscle volumes were measured during one session *pre* and *post* the resistance training program using freehand 3D ultrasound imaging to estimate muscle hypertrophy. One-repetition maximum (1RM) was also measured during one dedicated session *pre* and *post* the resistance training program to estimate strength transfer between the two exercises.

### Subjects

Thirty-six physically active (∼9h/week) sports science students participated in this study. The participants were familiar with resistance training, while none were following a specific hamstring-strengthening program. Inclusion criteria required participants to not be engaged in any specific lower limb training program and to have no history of lower limb injuries. Before the first experimental session, participants were randomly assigned to one of three groups: Control (CON, *n* = 12, age: 19.6 ± 1.1 years, height: 177.3 ± 7.7 cm, mass: 71.6 ± 10.6 kg), Nordic Hamstring Exercise (NHE, *n* = 12, age: 20.3 ± 1.5 years, height: 180.1 ± 9.1 cm, mass: 76.0 ± 11.1 kg), or Stiff-leg DeadLift (SDL, *n* = 12, age: 20.4 ± 1.4 years, height: 178.5 ± 7.1 cm, mass: 69.5 ± 9.3 kg). No participants dropped out the experimental protocol. Participants were instructed to maintain their usual physical activities, but to stop any lower limb resistance training. During an initial meeting, participants were informed about the nature, aims, and risks associated with the experimental and training procedures, and written consent was obtained. This trial (NCT06164249) was conducted in accordance with the Declaration of Helsinki and was approved by the ethics committee (No. 2021-A02993-38).

#### Dietary supplementation

In line with recommendations for protein intake during resistance training focused on hypertrophy (26), participants consumed a supplement on training days containing 22.0g of protein and 2.0g of carbohydrates (Whey Protein Bio, Alter Nutrition) under the examiner’s supervision.

### Procedures

#### Familiarization session

During the familiarization session, each participant was familiarized with both exercises. Since the Nordic hamstring often exceeds body weight as a supramaximal exercise, participants used a specific machine (Westside inverse curl pro; Watson, UK) equipped with a counterweight system that allowed off-loading the participants’ body weight and precisely quantify the load (Figure 1). After key execution criteria (detailed later) were presented and demonstrated by an experienced examiner, participants spent 10 minutes per exercise to (i) experience the full range of motion with a low resistance and (ii) perform 8 to 10 contractions at a moderate resistance while adhering to the execution criteria. If execution was validated by the examiner (otherwise, an additional set was performed), a progressive increase in load was performed over 3-4 sets to determine the maximal load for one repetition at a rating of perceived effort (RPE) of about eight (RPE 8). This maximal load performance was recorded and later used during the first one-repetition maximum (1RM) assessment session (*pre*).

**Figure 1.**
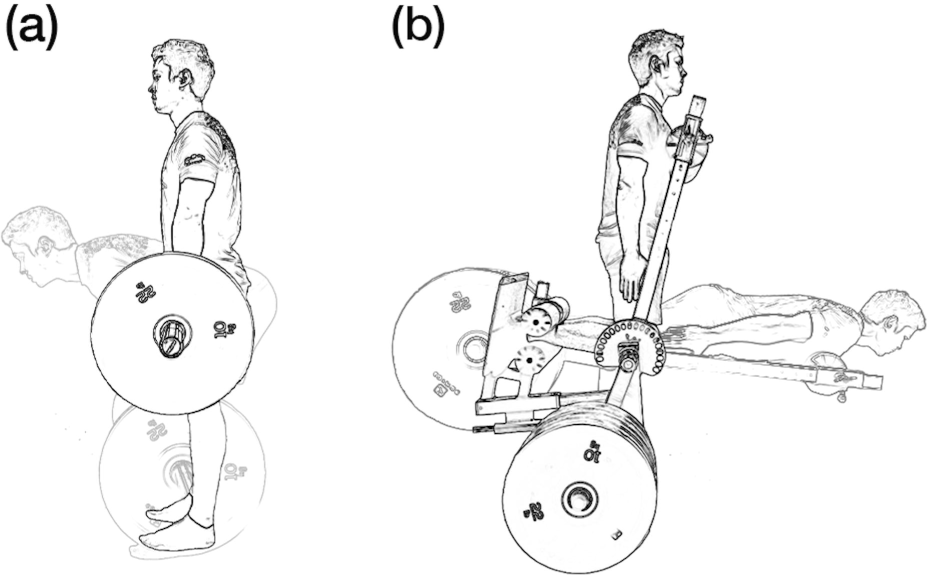
Representation of stiff-leg Deadlift (a) and Nordic hamstring exercise (b).

Nordic hamstring was performed from 90° knee flexion to full knee extension (thigh parallel to the floor) with a neutral spinal posture and complete hip extension (0°). Stiff-leg Deadlift was performed with a shoulder width stance, no lower limb rotation, and movement from 0° to 90° of hip flexion. Only slight knee unlocking was permitted, and a neutral spinal posture was maintained (Figure 1). For both exercises participants were asked to perform the eccentric phase over 3 seconds before executing the concentric phase with the intention of achieving maximal speed.

#### One-Repetition Maximal (1RM)

The 1RM was assessed for both exercises 4-10 days before and 4-7 days after the 9-week training program. To support the training session, the 1RM was also re-assessed at the start of the 10^th^ and 19^th^ training sessions. The 1RM evaluation session followed a standardized protocol to allow comparison across participants: participants completed a standardized warm-up consisting of 2 rounds of 10 body-weight single-leg bridge reps and an 8kg-loaded good morning exercise, with progressively increasing effort levels. This was followed by a specific warm-up (either Nordic hamstring or stiff-leg Deadlift) comprising 3 sets of 10, 6, and 1 repetitions performed at 30%, 60%, and 90% of the maximal load determined during the familiarization session, respectively. After the warm-up, a progressive load increase (minimum. 2.5 kg) was used to estimate the 1RM. If participants failed, a second trial was attempted after 3 minutes of rest. A valid 1RM required (i) a controlled three-second eccentric phase and (ii) compliance with the joint angles specified by the experimenter (see *Familiarization session*).

For Nordic hamstring, the lower the additional weight, the better the performance. Given the influence of individual biomechanics on Nordic hamstring force production, the 1RM torque (Nm) was calculated as follows:

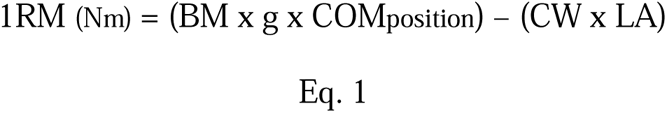

where BM is the participant’s body mass from Winter et al. (37), excluding (tibia and feet); g is the gravitational constant; COMposition is the estimated center of mass position of the system (body); CW is the additional counterweight; and LA is the lever arm of the machine counterweight.

For stiff-leg Deadlift, the 1 RM was the maximal load (kg) that could be lifted once over the range of motion (ROM). The 1RM for both the Nordic hamstring and stiff-leg Deadlift were tested in randomized order.

#### Training description

The training protocol took place at the “Movement – Interactions – Performance” laboratory of Nantes University (France) between February and April 2024. It consisted of 9 weeks of training for the NHE and SDL groups, with 3 sessions per week. The CON group continued their usual activities without any changes.

The NHE and SDL groups then began a progressive 27-session training protocol using a load set at 80% of their 1RM (33). As mentioned before, this load was adjusted every 3 weeks (5). Progressive changes in training volume (i.e., number of sets and repetitions) followed a pyramid structure (5) and are reported in Table 1. Execution criteria and participant’s safety were monitored by a certified trainer. The trainer was not involved in data collection or experimental data processing. Importantly, the training protocol was conducted using the exact same execution criteria as during the 1RM assessment.

**Table 1.**
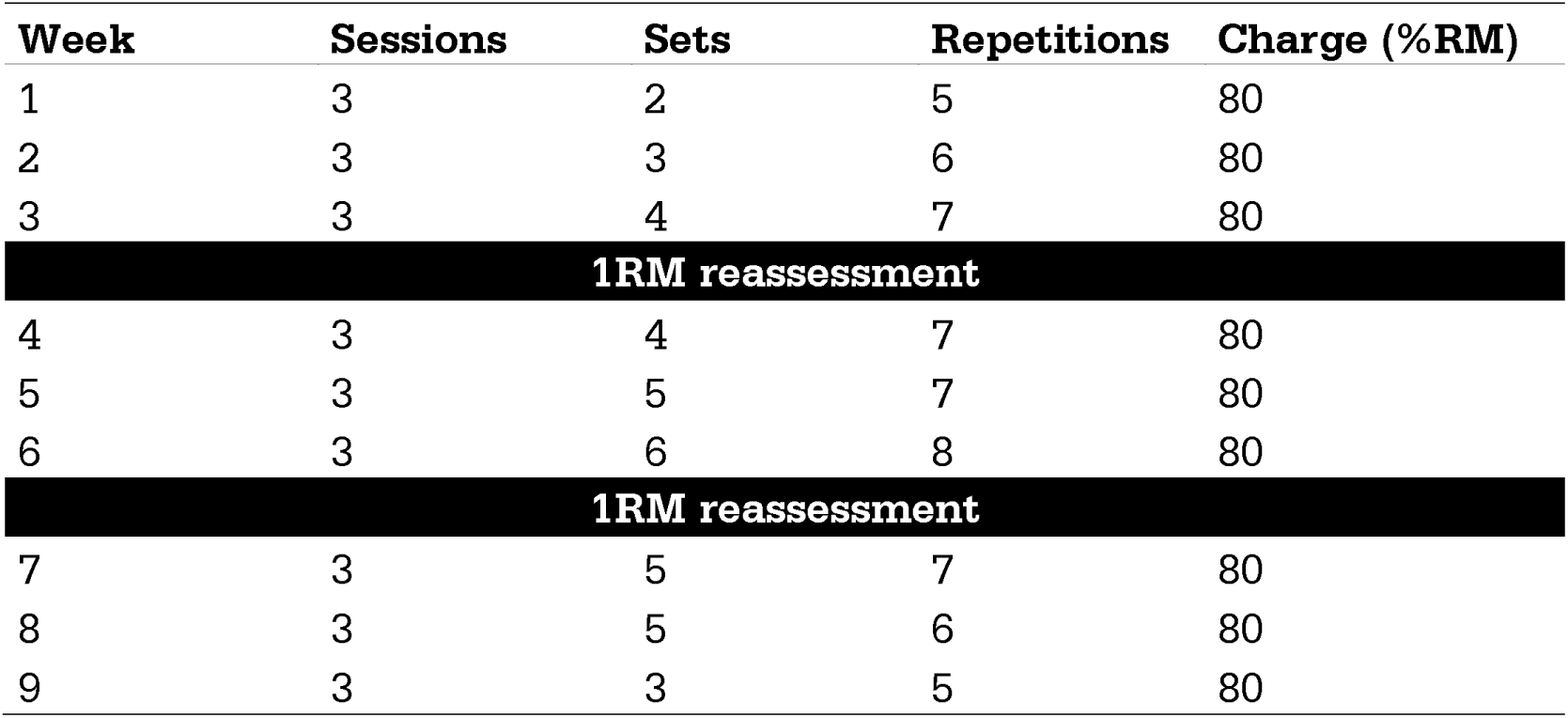
Training program for both Nordic hamstring and stiff-leg Deadlift exercises.

#### Resistance training monitoring

Participants monitoring during the 9-week training program was conducted using the HexFit application (Hexfit Inc, Quebec). This tool allowed participants to schedule laboratory appointments for experimental sessions and to book training slots for the NHE and SDL groups. Two participants (one from the NHE group and one from the SDL group) were permitted to train simultaneously, ensuring proper supervision by the strength and conditioning trainer. The application also enabled precise tracking of training loads throughout the 9 weeks. Additionally, it allowed the collection of data such as RPE, training-related comments, and fatigue levels.

#### Muscle volume assessment using 3D Ultrasound (3DUS) and hypertrophy estimation

The freehand 3DUS method used in the present study has been validated for the hamstrings (12). Participants laid prone on a massage table while ultrasound images of each hamstring head were captured using an ultrasound scanner (Aixplorer version 12.3, SuperSonic Imagine, Aix-en-Provence, France) with a 10-2 MHz transducer (40 mm field of view: Vermon, Tours, France). Image depth and brightness were individually adjusted for each participant.

The transducer was fitted with a 3D-printed rigid body containing four reflective markers. An optoelectric motion capture system with ten cameras (Optitrack Flex 13, NaturalPoint, USA), recording at 120 Hz, tracked the transducer’s position. Ultrasound images and transducer position data were recorded using the open-source software 3D Slicer (slicer.org; v.5.6.2; Perth, Australia). To ensure measurement reproducibility, the same examiner (T.M.) conducted all pre- and post-training volume recordings. Furthermore, for each measurement, the examiner was blinded to the participant group assignments. A gel pad was used to minimize compression effects, and no pressure was applied to the transducer during imaging (12,35).

Once data collection was complete, all muscle volumes were reconstructed and manually segmented using 3D Slicer by the same blinded examiner for consistency. Manual segmentations were conducted every 6.0mm at the hamstring insertions and every 10.0mm in the muscle belly regions, representing approximately 30 to 50 slices per muscle. Hypertrophy of each individual muscle was considered as the relative changes in muscle volume between *pre* and *post.* Whole hamstring hypertrophy was calculated as the averaged hypertrophy of the three hamstring heads (i.e., SM, ST, and BF).

#### Muscle activation

Myoelectrical activity was recorded using surface EMG *pre* and *post* training during the 1 RM. EMG electrodes were placed over the SM, ST, and BF muscles. Before electrode placement, the skin was shaved and cleaned with alcohol. Wireless surface electrodes (Cometa Pico, Cometa, Milan, Italy, interelectrode distance = 25 mm) were attached to the skin with double-sided tape (inter-sensor distance = 1 cm). Electrode placement was verified with B-mode ultrasound (6) to ensure alignment with the muscle fascicle plane and avoidance of neighboring muscle borders. To minimize crosstalk, EMG electrodes were placed on the midline of the most prominent superficial muscle belly of each hamstring head. Based on hamstring morphology (34), the electrode positions for BF and SM were slightly more distal than for ST. Given the regional variation in EMG amplitude (15,16), two electrodes were placed on each muscle: one at the proximal-middle and one at the distal-middle location. EMG signals were digitized at 2000 Hz and band-pass filtered (30–300 Hz) using a 2^nd^-order Butterworth filter. The signals were then rectified and low-pass filtered at 6 Hz to generate individual EMG envelops (17). For EMG normalization, two maximal voluntary contractions (MVCs) were performed at three different knee flexion angles (20°, 45° and 80°; 0° = knee fully extended) with a fixed 90° of hip flexion (0° = full hip extension) (27). The peak value of each EMG envelops over the six MVCs was used as the maximal reference value for normalizing EMG signals to obtain an estimate of muscle activation. Kinematic data (MTw Awinda, Xsens North America Inc., Culver City, USA) was used to determine the onset and offset of each repetition of an eccentric contraction. Muscle activation was measured during the ROM as follows: from 90° to 0° of knee flexion for Nordic hamstring, and from 0° to 90° of hip flexion for stiff-leg Deadlift. Activation was expressed as percentage of the ROM for each exercise.

### Statistical Analysis

The data distribution was assessed using the Shapiro-Wilk test. Outliers were detected using Grubbs’ test (14), resulting in the exclusion of one participant of the CON group from the analyses. Absolute (kg and Nm) and relative (%) strength gains (1RM) were calculated for all participants.

The effect of the 9-week training program on 1RM in the trained exercise (Nordic hamstring or stiff-leg Deadlift) was evaluated using two independent Student’s T-tests to compare relative strength gains between control and trained groups (CON vs. NHE, CON vs. SDL). To assess differences in strength transfer between the NHE and SDL groups, a Student’s T-test was used to compare relative 1RM changes in the non-trained exercise.

A linear mixed-effects model was used to evaluate differences in muscle hypertrophy (%) between groups (CON, NHE, and SDL) and muscles (SM, ST, and BF). Group and Muscle were modeled as fixed effects, along with their interaction (Group x Muscle) to examine whether hypertrophy responses differed across muscles depending on the group. To account for inter-subject variability, subject was included as a random effect.

To compare EMG patterns with high temporal resolution, all statistical analyses were performed using Statistical Parametric Mapping (SPM 30, v0.4, www.spm1d.org) in Matlab® (Matlab; The Mathworks, Nathick, MA, USA). Family-wise type I error rate was set at 0.05. Two three-way repeated-measures ANOVA was used to determine whether time (*pre* and *post*), muscle (SM, ST, and BF), group (NHE vs. CON; SDL vs. CON), and their interactions altered EMG signals recorded during the non-trained exercise. In case of an interaction at any timepoint across the range of motion, locations of the differences were tested using paired-sample t-tests with Bonferroni correction. SPM technical details are described elsewhere (1,11).

Finally, correlations were performed (by group) to determine the relationship between whole hypertrophy, selective hypertrophy and changes in 1RM for the non-trained exercise. Correlation coefficients were interpreted as negligible, small, moderate, or large at <0.10, 0.11-0.30, 0.31-0.50, and >0.50, respectively (9).

Effect sizes were also calculated using Cohen’s d (9) and interpreted as *small*, *medium* and *large* at <0.20, 0.21-0.50 and 0.51-0.8, respectively. All statistical analyses and data visualizations in this study were performed using R (R Foundation for Statistical Computing, v.4.4.0, Vienna, Austria) and Prism (GraphPad Prism, v.10.1.1, Boston, Massachusetts USA) software. Data tables and statistical results are available as supplementary materials.

## RESULTS

Training session attendance was 96.6 ± 3.7% for the NHE group and 94.8 ± 4.0% for the SDL group. Table 2 shows the 1RM results for each group in both trained and non-trained (strength transfer) exercises. Individual progressions in Nordic hamstring and stiff-leg Deadlift exercises for the NHE, SDL and CON groups are presented in Figure 2.

**Figure 2.**
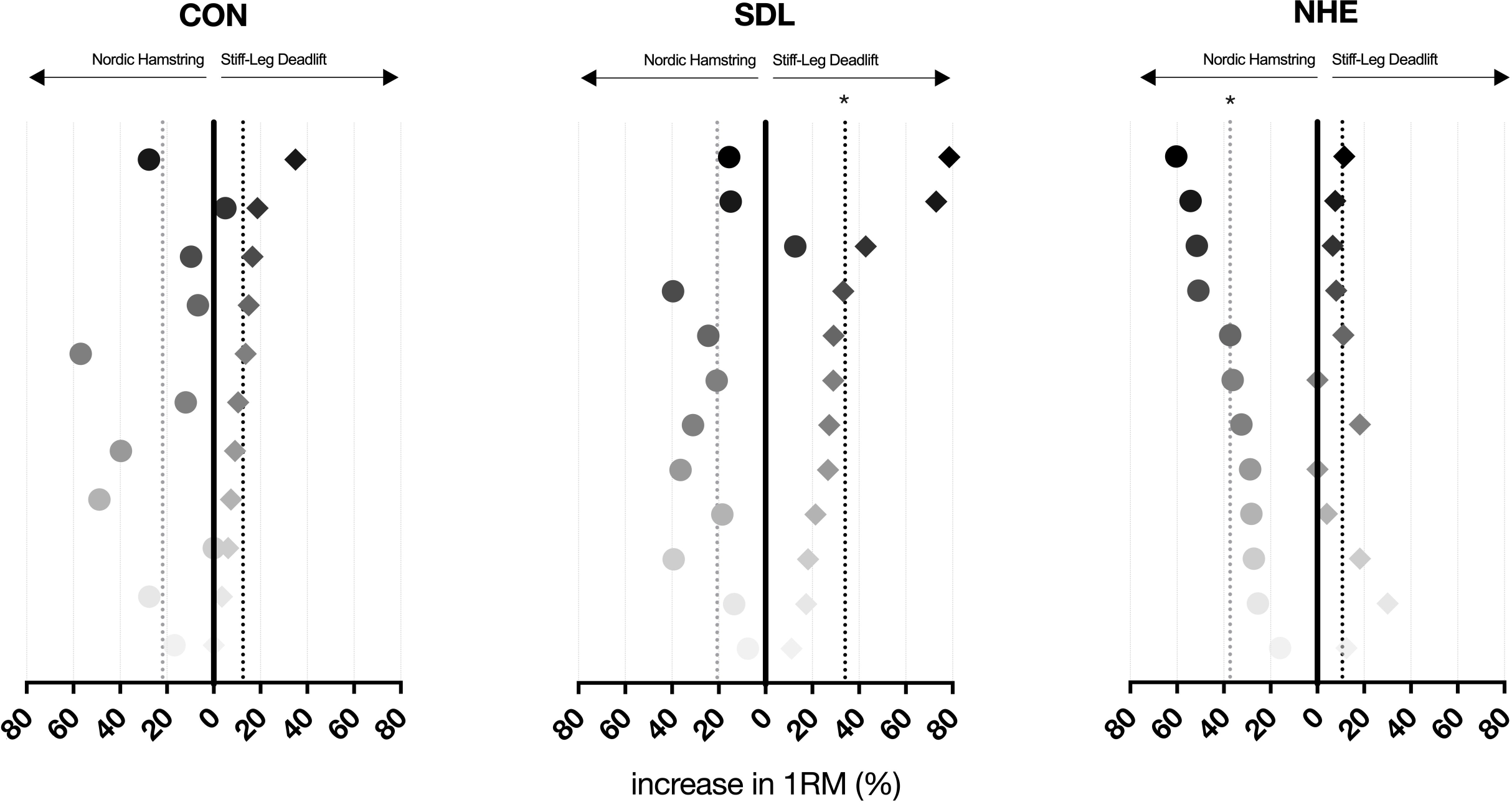
Relative increases in 1RM for each group (i.e., CON, SDL, and NHE), for each exercise and participant. The data for the Nordic hamstring exercise trends to the left, while the data for the stiff-leg Deadlift exercise trends to the right. The mean increases for the Nordic hamstring exercise are indicated by gray vertical dotted lines, whereas the mean increases for the stiff-leg Deadlift exercise are marked by black vertical dotted lines. *: significantly different from the CON group.

**Table 2.** Changes in 1RM for both exercises and Control (CON), Nordic Hamstring Exercise (NHE) and Stiff-Leg Deadlift (SDL) groups. Pre: 1RM before the training program; post: 1RM after the training program.

| Group | stiff-leg Deadlift |  | Nordic hamstring exercise |  |
| --- | --- | --- | --- | --- |
|  | pre | post | pre | post |
| **NHE (n = 12)** | 125.4 ± 30.0 kg | 138.1 ± 30.6 kg | 201.0 ± 60.2 Nm | 274.6 ± 81.6 Nm |
| **SDL (n = 12)** | 103.5 ± 22.9 kg | 136.0 ± 23.0 kg | 198.2 ± 80.5 Nm | 232.0 ± 85.3 Nm |
| **CON (n = 11)** | 115.5 ± 26.5 kg | 128.2 ± 22.9 kg | 183.6 ± 53.2 Nm | 221.3 ± 61.4 Nm |

### Muscle hypertrophy

The linear mixed-effects model revealed a significant group muscle interaction effect on changes in muscle hypertrophy (p < 0.001) as well as a significant group effect (p = 0.004). Post-hoc tests showed that both the NHE and SDL groups experienced significant overall hamstring hypertrophy compared to the control group (p < 0.001 and p = 0.044, respectively). No significant difference in overall hypertrophy was observed between the NHE and SDL groups (p > 0.05).

Regarding muscle-specific hypertrophy, the NHE group exhibited significant hypertrophy of the ST (24.4 ± 10.8%) compared to both the CON group (1.0 ± 6.2%; p < 0.001) and the SDL group (5.5 ± 9.3%; p < 0.001). The SDL group, in contrast, showed significant hypertrophy of the SM (11.2 ± 12.7%) compared to the CON group (1.9 ± 4.3%; p = 0.02), but not compared to the NHE group (4.6 ± 5.8%; p = 0.24). For the BF, no significant differences in hypertrophy were observed between any of the groups (p ≥ 0.39).

### Relative strength gains in the trained exercise

In the Nordic hamstring exercise, the NHE group showed a significantly larger increase in 1RM compared to the CON group (mean difference = 18.8%, 95% Confidence Interval (CI): 3.0-34.5%, p = 0.02, d = 0.8). On average, participants in the NHE group exhibited a 37.4 ± 13.8% increase on 1RM after 9 weeks of training (Figure 2). In the stiff-leg Deadlift exercise, the SDL group showed a significantly larger increase in 1RM compared to the CON group (mean difference = 21.6%, 95% CI: 7.2-36.1%, p = 0.01, d = 1.0). On average, participants in the SDL group exhibited a 34.0 ± 21.2% increase in 1RM.

### Relative strength gains in the non-trained exercise (strength transfer)

The relative increase in 1RM in the non-trained exercise was 10.7 ± 8.5% and 20.7 ± 15.0% in the NHE group and the SDL group, respectively (Figure 2). Additionally, no significant difference was found between the CON group and the respective progressions of the NHE group in stiff-leg Deadlift (mean difference = 1.7%, 95% CI -9.5-6.1%, p = 0.65; d = 0.2) and the SDL group in Nordic hamstring (mean difference = -1.2%, 95% CI -14.1-16.5%, p = 0.87; d = 0.1). No significant difference in strength gains in the non-trained exercise was observed between the NHE and SDL groups (mean difference = 10.1%, 95% CI 0.3-20.4%, p = 0.06; d = 0.8).

### Muscle activations in the non-trained exercise

The muscle activation patterns are depicted in Figure 3. SPM analyses revealed neither a time, group nor a time muscle group (all p ≥ 0.05) effect on muscle activations during the non-trained exercises. A main effect of muscle was observed for both Nordic hamstring (p < 0.001) and stiff-leg Deadlift exercises (p < 0.001).

**Figure 3.**
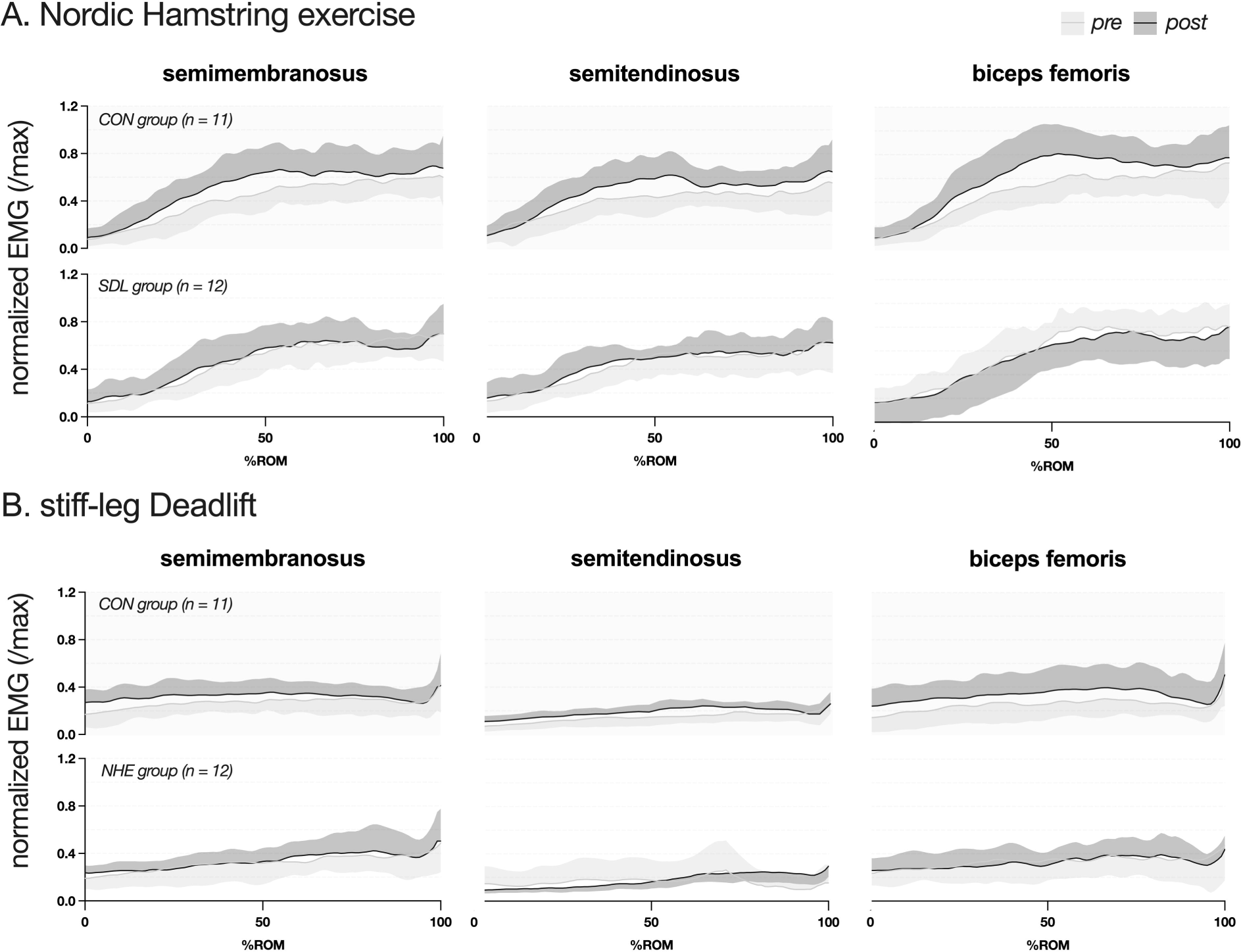
Normalized EMG for each hamstring head pre- and post-training measurements for non-trained exercise and for both exercises for CON group. Pre-training are represented by gray lines and areas, while post-training EMG data are represented by black lines and areas. The data are presented as a function of the range of motion (from 0 to 100%) during the eccentric phase of the movement. Panel A: data for the SDL and CON groups in Nordic hamstring. Panel B: data for the NHE and CON groups in stiff-leg Deadlift.

#### Correlation between strength transfer and muscle hypertrophy

For both the NHE and SDL groups, a non-significant correlation was observed between the increase in total hamstring hypertrophy and the strength transfer to non-trained exercise (r ≤ 0.23; p = 0.52). The relationships between selective hypertrophy of the SM, ST, and BF and strength transfer to the non-trained exercise, depicted in Figure 4, yielded similar results. For the SDL group, *negligible* to *small and* non-significant correlations (r ≤ 0.27; p ≥ 0.39) were found between SM, ST, and BF hypertrophy and strength transfer to Nordic hamstring exercise (bottom panels in Figure 4). For the NHE group, *small* and *moderate* non-significant correlations (r ≤ 0.40; p ≥ 0.19) were found for SM and ST (top panels in Figure 4), while a *large* and significant positive correlation was observed between BF hypertrophy and strength transfer to stiff-leg Deadlift exercise (r = 0.60; p = 0.04).

**Figure 4.**
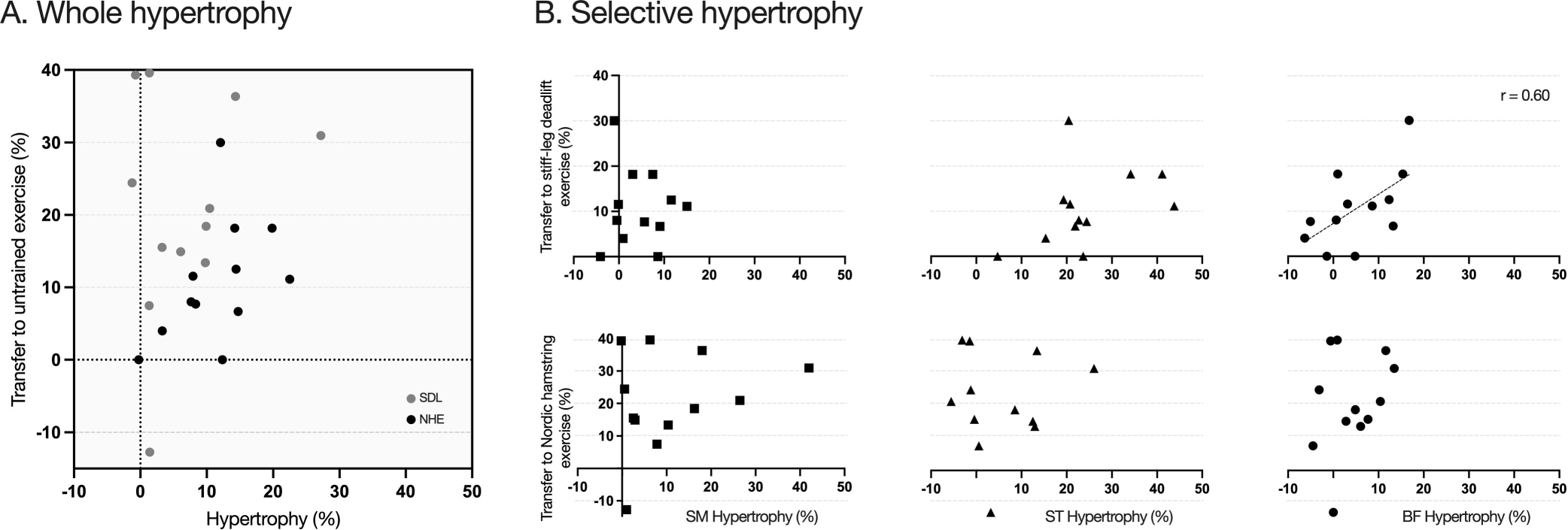
Relationship between strength transfer and hypertrophy. The Panel A represents the relationship between whole hypertrophy and strength transfer in the untrained exercise for every participant. Data for the SDL group are represented by gray dots while data for the NHE group are represented by black dots. The Panel B represents the relationship between strength transfer in untrained exercise and selective hypertrophy for the NHE and SLD groups. Data of NHE group are presented in top Panel. Data of SDL are presented in bottom Panel.

## DISCUSSION

We conducted a blinded randomized controlled trial over 9 weeks to examine the effects of Nordic hamstring and stiff-leg Deadlift exercises on strength gains in non-trained exercises, and (i) whole and (ii) selective hamstring hypertrophy, and conclude on the possible correlation between muscle hypertrophy and strength transfer. In presence of a significant whole hamstring hypertrophy in NHE (11.4 ± 6.5%) and SDL (7.0 ± 8.1%) groups, they did not exhibit larger strength transfer compared to CON. According to our hypothesis, NHE group exhibited a selective hypertrophy of the ST (24.4 ± 10.8%) while SDL group exhibited a selective hypertrophy of the SM (11.2 ± 12.7%). However, no difference in strength transfer was observed between NHE and SDL groups. Finally, no significant correlations were found between both whole and selective hamstring hypertrophy and strength transfer. We concluded a minimal role of hamstring hypertrophy in strength transfer between Nordic hamstring and stiff-leg Deadlift in resistance-untrained individuals.

Consistent with prior research, Nordic hamstring curl-based resistance training effectively improved maximal knee flexion strength (2,3,5,7,22). Previous studies, such as Andrews et al. (3), reported a ∼40% increase in eccentric knee flexion torque after 9 weeks of Nordic hamstring training, which is in line with the 37.4 ± 13.8% increase in 1RM observed here for the NHE group after 27 training sessions. In contrast, stiff-leg Deadlift training has been less explored, although Kawama et al. (18) reported 20-26% increases in maximal isometric knee flexion torque. However, this study used lower loads (60% of body mass), maximal range of motion to focus on flexibility and hamstring structural adaptations and did not report results in terms of 1RM. Our study, which employed different loads (∼80% 1RM), also observed a large increase in strength in the trained stiff-leg Deadlift exercise (34.0 ± 21.2% increase in 1RM for the SDL group; Figure 2).

Both the NHE and SDL groups exhibited significant and similar whole hamstring hypertrophy compared to the CON group (11.4 ± 6.5% and 7.0 ± 8.1%, respectively, vs. 1.2 ± 3.6%). Albeit no existing Nordic hamstring-stiff-leg deadlift comparison in the literature, these results align with Bourne et al. (5), who reported 15-16% hamstring hypertrophy for the Nordic hamstring, with no difference compared to a 45° hip extension exercise. The similar participants training status (non-resistance trained) combined with the similar sets and repetitions range (Table 1) likely explain the accordance with our study. According to our second hypothesis, the NHE and SDL groups exhibited significantly greater selective ST and SM hypertrophy, respectively. Although this is the first study comparing directly these two exercises, this result is in accordance with the literature (5,18). Inter-individual variability of the distribution of hypertrophy was notable, with some participants from the NHE group notably showing 80-100% of the total hamstring hypertrophy in ST, while another from the same group showed only 44%. Similar results were observed in the SDL group, as one participant showed 76% of the total hamstring hypertrophy in SM, while it represents 24% for another participant from the same group. The interindividual difference in the distribution of the hamstring hypertrophy has been already reported in Frouin et al. (13). The identification of the origin and the consequences of this phenomenon requires further investigations.

Carmichael et al. (8) reported that a 6-week eccentric and isometric hip extension exercise intervention induced whole hamstring hypertrophy of about 10% and 8.5%, respectively. Strength transfer to knee flexion performance followed specificity principle with significant gains in eccentric condition (12.0% and 8.4% at moderate and fast speed, respectively) and non-significant gains in isometric condition for the eccentric group, with reciprocal results for the isometric group (10.3% in maximal isometric knee flexion strength). This a crucial result, as it suggests that 10% and 8.5% of hypertrophy of both groups do not enhance the maximal strength in the knee flexion performance in the non-trained contraction regimen. Although we reported substantial increases in 1RM in the non-trained exercise (10.7% and 20.7% for NHE and SDL group, respectively), these changes were not different compared to CON (Figure 2). This means that the significant increase in hamstring volume in both the SDL (7.0 ± 8.1%) and NHE (11.4 ± 6.5%) groups did not contribute to enhance strength transfer. Also, the greater selective SM hypertrophy induced in SDL group, which exhibit large torque generating capacities (4), did not led to a larger strength transfer to Nordic hamstring (non-trained exercise), which was counterintuitive to our initial hypothesis. Overall, no significant relationship was found between strength transfer and whole hypertrophy (all r values ≤ 0.23), and selective hypertrophy of each hamstring head (all r values ≤ 0.4), except for the BF in stiff-leg Deadlift (r = 0.6; Figure 4). This latter *large* correlation needs to be interpreted with caution as the magnitude of hypertrophy on BF was about 6.3% on average, with 7/12 participants exhibiting less the 8% of hypertrophy. This provides the first impetus that hypertrophy plays a minimal role in strength transfer in non-resistance trained individuals. Together with the absence of changes in muscle activation between *pre* and *post* resistance training for any group (Figure 3), this helps us conclude about the major role of technical factor (i.e., the participant’s ability to produce efficient force) in the strength transfer observed in NHE and SDL groups, as well as the strength gains in the CON group.

Three main hypotheses may be formulated to explain the minor role of hypertrophy in strength transfer. First, the present study has shown that hamstring muscle activation is submaximal during the 1RM of Nordic hamstring and stiff-leg Deadlift exercise (figure 2). It is likely that the fibers exhibiting hypertrophy were not all recruited during 1RM reducing the hypertrophy-related strength transfer. Second, Hegyi et al. (15) showed a region-dependent hamstring electrical activity in Nordic hamstring exercise and stiff-leg Deadlift with proximo-distal patterns depending on muscle and exercises. If the Nordic hamstring exercise induced hypertrophy to the distal region of the ST (21), the low activation of this region during stiff-leg Deadlift would limit the strength transfer. In a similar manner, musculoskeletal modelling revealed that the contribution in total force was limited for ST and SM in stiff-leg Deadlift and Nordic hamstring exercises, respectively (36). This limits the role of selective hypertrophy in strength transfer between these two specific exercises. Third, the strength transfer associated with muscle hypertrophy requires that hamstring strength represent a limiting factor of exercise performance. This is a large assumption as many other factors (non-technical) may have limited performance in stiff-leg Deadlift, such as grip (29), spinal extension (30) and gluteal (28) maximal strength. This is particularly true in bilateral condition with a high relative load compared to Nordic hamstring curl performed on a guided machine. Although speculative, it is likely that the strengthening of such “accessory” or “non-primary” muscles in SDL group may contribute to strength transfer in Nordic hamstring exercise. In line with this latter hypothesis, we found a *large* (non-significant) greater strength transfer from SDL group toward Nordic hamstring exercise than those from NHE group toward stiff-leg Deadlift exercise. Note that the opposite trend observed for CON group reinforces this hypothesis. This suggests that strength transfer between Nordic hamstring and stiff-leg deadlift exercise is mediated by other factors (non-technical), the magnitude of which depends on the exercise.

### Limitations and statistical considerations

The interpretation of statistical results in this study should consider certain limitations. The small sample size (36 subjects across three groups) likely reduced the statistical power to detect subtle relationships between hypertrophy and strength transfer, increasing the likelihood of Type II errors (e.g., concluding p > 0.05 and incorrectly interpreting absence of significant correlation while a moderate linear relationship of r = 0.41 between hypertrophy and strength transfer could exist). This issue is compounded by large inter-subject variability observed in both hypertrophy and strength transfer responses, and captured by the mixed-effect model in the hypertrophy versus group-muscle analysis.

Such variability increases noise in the data, widening confidence intervals and making it difficult to detect consistent trends. Together, these factors underscore that larger sample sizes are needed to account for individual differences, improve the reliability of the statistical estimates, and consolidate the conclusions drawn in this study.

### Future directions

Future studies should explore whether hypertrophy accelerates strength gains during a subsequent training block focused on the previously untrained exercise from the first block. In line with the micro-dosing principle, it would be valuable to design a mixed program combining a high training volume for one exercise (designated as the trained exercise) with a minimal effective dose for another (designated as the untrained exercise). This approach could help us to determine whether muscle hypertrophy has a greater impact on strength transfer when the technical dimension is minimized. This question should also be examined in resistance-trained individuals. Such an approach would offer deeper insights into the role of muscle hypertrophy in performance (25) and its broader implications for rehabilitation and training strategies.

## PRACTICAL APPLICATIONS

Coaches and practitioners should include both Nordic hamstring curl and stiff-leg Deadlift exercises in the same training block to maximize strength gains in both knee flexion-oriented and the hip extension-oriented tasks. From the results of our study, both whole and selective hypertrophy induced by Nordic hamstring and stiff-leg Deadlift exercises did not affect the magnitude of strength transfer in resistance-untrained participants. The current study suggest that strength transfer was also mediated by other non-technical factors (e.g., strengthening of non-primary muscles), the magnitude of which depends of the exercise. Future research needs to determine the influence of resistance training status on hypertrophy-induced strength transfer and its putative optimization through minimal effective dose during a block periodization.

## Data Availability

All data produced in the present study are available upon reasonable request to the authors

## ACKNOWLEDGMENTS

The authors thank all subjects for their engagement in the study, Romain MAHE for the supervision of the resistance training program, and the *Region Pays de la Loire* for the grant *Étoiles Montantes* conceded to Lilian LACOURPAILLE. The authors declare that they have no conflict of interest regarding the publication of this paper.

## Notes

### Competing Interest Statement

The authors have declared no competing interest.

### Clinical Trial

NCT06164249

### Funding Statement

Region Pays de la Loire for the grant etoiles Montantes conceded to Lilian LACOURPAILLE

### Author Declarations

Ethics Committed Paris IV gave ethical approval for this work 2021A0299338

